# Social and Cardiovascular Risk Factors as Predictors of the Progression from Mild Cognitive Impairment to Dementia in a Large EHR Database

**DOI:** 10.64898/2026.03.02.26347451

**Authors:** Silvia Miramontes, Erin L. Ferguson, Scott Zimmerman, Evan Phelps, Boris Oskotsky, Tomiko T. Oskotsky, John A. Capra, Elena Tsoy, Marina Sirota, M. Maria Glymour

## Abstract

**Background and Objectives:** Progression from mild cognitive impairment (MCI) to Alzheimer’s Disease and Related Dementias (AD/ADRD) varies widely across individuals, yet the mechanisms underlying this heterogeneity remain unclear. Identifying clinical and social determinants influencing this transition could enable earlier intervention. While cardiovascular and social risk factors are established contributors to dementia incidence, their role in progression from MCI to dementia may differ. Few studies using real world clinical data have evaluated these potential determinants of MCI progression.

**Methods:** Using electronic health records (EHR) from patients with incident MCI at UCSF Health (2010-2024), we evaluated cardiovascular (blood pressure [BP], body mass index [BMI], and type II diabetes) and social (marital status, language preference, race/ethnicity, and neighborhood disadvantage) risk factors for rate of progression from MCI to AD/ADRD. Covariate-adjusted Cox proportional hazards models estimated hazard ratios for incident AD/ADRD, with evaluation of interactions by sex.

**Results:** Among 6,529 patients, higher systolic BP was associated with AD/ADRD incidence (HR per 10 mmHg: 1.09, 95% CI: 1.05–1.14). BMI was inversely associated with incidence in both males (HR: 0.94; 95% CI: 0.92–0.97) and females (HR:0.98; 95% CI: 0.96–0.99). Compared to married individuals, widowed patients had a higher hazard of progression (HR: 1.15; 95% CI: 1.00–1.32). Spanish-speaking (HR: 1.38; 95% CI: 1.04–1.81), Chinese-speaking (HR: 1.19; 95% CI: 1.00–1.42), and “Other non-English” speaking patients (HR:1.24; 95% CI: 1.03–1.51) had a higher hazard of progression compared to English speakers. Latinx (HR:1.22; 95% CI: 1.01–1.48) and Asian patients (HR:1.14, 95% CI: 1.00-1.30; p=0.04) also had higher hazards of progression compared to White patients. Neighborhood disadvantage was not significantly associated with disease progression.

**Discussion:** Cardiovascular and social factors independently influence dementia progression, with some sex-specific patterns. Integrating clinical and social indicators highlights the potential of EHR data to identify high-risk patients earlier in the care continuum and support equitable dementia prevention.

## BACKGROUND

Each year, approximately 10–15% of individuals with mild cognitive impairment (MCI) progress to Alzheimer’s disease (AD) or Alzheimer’s Disease and related dementias (AD/ADRD) [1–3]. MCI is characterized by measurable cognitive decline that does not yet interfere substantially with daily functioning [4], representing a critical, potentially actionable phase in the dementia continuum. Not all individuals with MCI progress to dementia and time to transition to dementia varies substantially, introducing heterogeneity that complicates prognosis and care planning. Identifying clinical and social risk factors that accelerate this transition could enable earlier intervention, targeted monitoring, and more equitable care strategies. Yet most prior studies have emphasized biological or biomarker-based predictors of transition from MCI to dementia, leaving clinical, social, and vascular determinants – key drivers of dementia risk in the general population – largely underexamined.

Many predictors of cognitive decline exhibit paradoxical or disease-stage-specific effects during the MCI to AD/ADRD transition. For example, higher educational attainment is generally associated with lower dementia incidence [5], yet faster functional decline once MCI or dementia is diagnosed, potentially reflecting delayed recognition of impairment due to cognitive reserve [6]. Similarly, vascular and metabolic risk factors such as hypertension, body mass index (BMI), and diabetes have been widely studied but the evidence remains inconsistent. Diabetes and metabolic syndrome are robustly associated with greater risk of progression from MCI to dementia [6–12], results for hypertension have ranged from protective to harmful [7,11,13–15]. Much of this inconsistency likely reflects design limitations: most studies do not begin follow-up at incident MCI diagnosis, leaving the initial time at risk uncertain; many of the prior studies rely on small, homogenous, or highly selected samples. The Alzheimer’s Disease Neuroimaging Initiative (ADNI) is a commonly used data source, but ADNI is not only a highly selected sample, it enrolls prevalent rather than newly diagnosed MCI cases. Beginning follow-up among prevalent MCI cases obscures the natural course of progression and introduces selection and survival biases. These limitations hinder comparability across existing studies and contribute to inconsistent conclusions regarding the role of vascular risk in the MCI-to-dementia transition.

Efforts to improve risk prediction have leaned on narrow, biomarker-focused models that are difficult to translate into routine clinical care. Predictive modeling studies have largely been derived from ADNI [6,16–20] or other specialized cohorts [10,16,21–24], prioritizing imaging and cerebrospinal fluid measures that are not available in primary care settings. While these data provide valuable physiological insights, their limited accessibility restricts generalizability to real-world-clinical settings, where most patients with MCI are first evaluated and where early identification of high-risk individuals could meaningfully alter the patient’s trajectory of care. Further, Black and Hispanic individuals are substantially less likely to undergo advanced biomarker testing due to disparities in specialty access and referral patterns [25,26], limiting the public health applicability of biomarker-based models in diverse real-world populations.

Recently published expert panels emphasize biomarkers as the strongest predictors of dementia progression [27], but this may reflect the paucity of research evaluating clinical and social measures available in typical clinical settings. Given these limitations, routinely collected measures recorded during primary care encounters, such as blood pressure, body mass index, language preference, and marital status, could provide early signals of elevated risk. Although social and structural determinants of health have been associated with dementia incidence [28], they remain understudied in the context of MCI progression, partly because they are inconsistently captured in electronic health records (EHRs).

In this study, we use EHRs from 6,529 patients with incident MCI seen at a large San Francisco Bay Area hospital system to evaluate clinical and social risk factors for progression to AD/ADRD. We integrate repeated longitudinal clinical measures with geospatial indicators of neighborhood context to assess how place-based social determinants of health contribute to differential risk of progression. Our combination of structured EHR data and area-level social data provides a scalable framework to quantify clinical and social influences on risk of progression, supporting earlier and more equitable identification of patients at elevated risk in real-world primary care settings.

## METHODS

This study was approved by the University of California, San Francisco (UCSF) Institutional Review board (IRB # 20-32422). The requirement for informed consent was waived due to the retrospective analysis of existing economic health record data.

### Study Population and Study Design

We identified 6,529 patients aged 50 years and older who received a clinical diagnosis of mild cognitive impairment (MCI) at UCSF between 2010 and 2024, in structured electronic health records (EHR) from specialty and primary care settings. Diagnosis of MCI and Alzheimer’s Disease or Alzheimer’s Disease and Related Dementias (AD/ADRD) were defined using International Classification of Diseases (ICD) codes (ICD-9 and ICD-10) recorded in the EHR. Specific codes used to identify MCI and AD/ADRD are listed in **eTables 1** and **2**, respectively.

To enhance diagnostic specificity, we restricted the identification of MCI diagnoses to encounters occurring within dementia specialty clinics (e.g., memory clinics, geriatrics) and internal medicine or primary care settings. These settings are more likely to include clinically valid cognitive assessments and represent the most likely locations where patients with suspected cognitive decline would receive a formal diagnostic evaluation. The primary outcome was time to incident AD/ADRD, with follow-up beginning at the date of first qualifying MCI diagnosis and ending at the earliest of: AD/ADRD diagnosis, death, or last recorded clinical encounter (**Figure 1B**).

**Figure 1.**
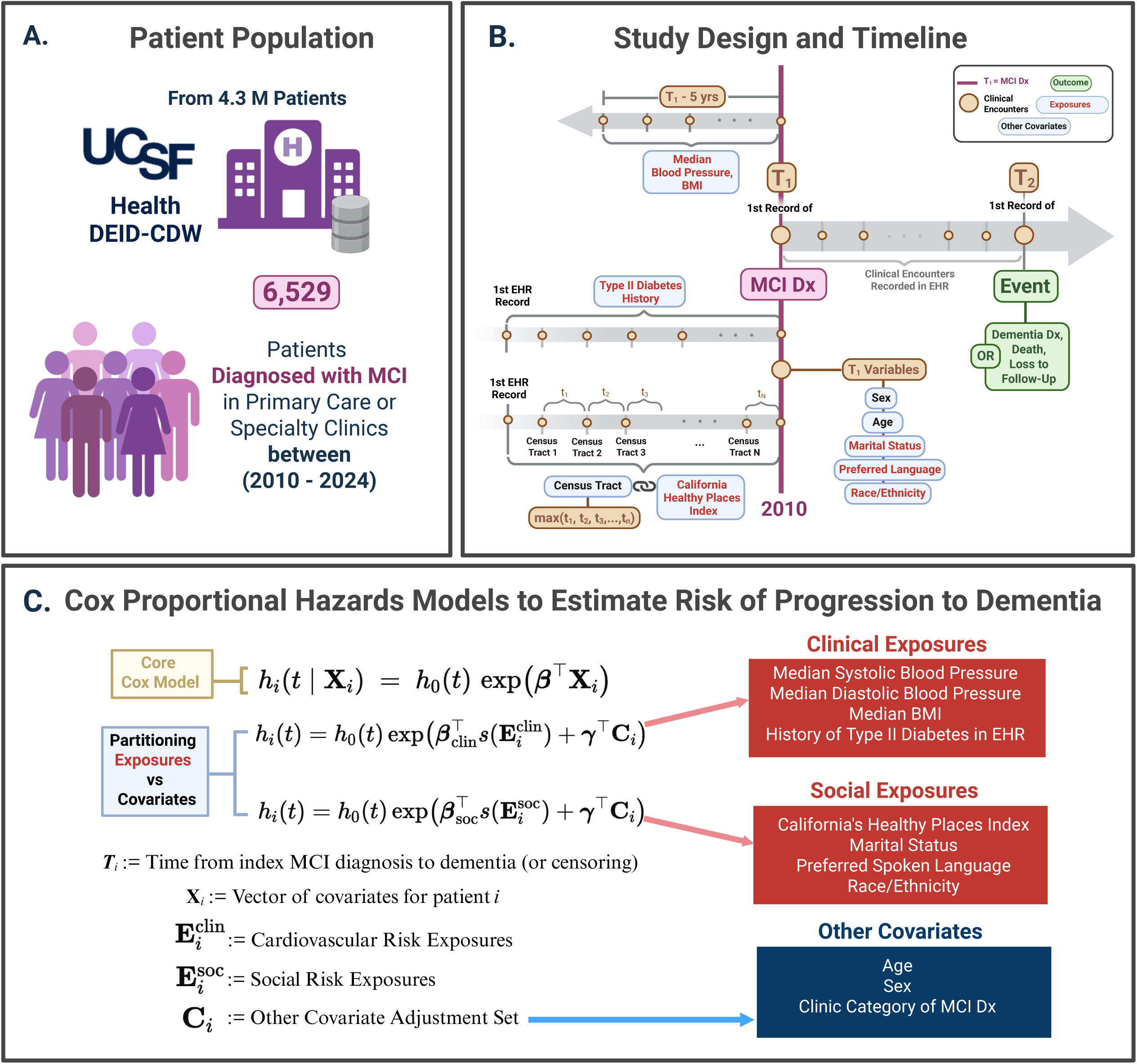
Study population, design, and modeling framework. A. From the UCSF Health de-identified clinical data warehouse (∼4.3M patients), we identified 6,529 adults with incident mild cognitive impairment (MCI) diagnosed in primary care or specialty clinics between 2010–2024. B. Timeline anchored at index MCI diagnosis T1. Clinical exposures were defined prior to T1: median systolic/diastolic blood pressure and BMI over the preceding 5 years; history of type 2 diabetes from any pre-T1 record. Social exposures were assigned using the California Healthy Places Index (HPI) for the longest-resided census tract prior to T1. Baseline covariates (age, sex, marital status, preferred language, race/ethnicity) were taken at T1. Follow-up continued from T1 to the earliest of dementia diagnosis (event), death, or last encounter (censoring). C. Cox proportional hazards models estimated the hazard of progression from MCI to dementia, with separate adjusted models for clinical/cardiovascular exposures and for social exposures (and an optional combined model). Continuous predictors may be modeled with spline terms; models adjust for age, sex, and clinic category of the MCI diagnosis.

### Exposure Variables: Social and Clinical Risk Factors

Social risk factors considered in this study included neighborhood disadvantage, marital status, racial/ethnic identity, and preferred spoken language. Neighborhood disadvantage was measured using the California Healthy Places Index (HPI) [29], a composite indicator of socioeconomic and environmental opportunity developed by the Public Health Alliance of Southern California [30]. We linked HPI scores (range: 1-100, higher values indicating greater neighborhood resources) to the census tract of each patients’ longest residence prior to MCI diagnosis based on EHR residential history, capturing long-term neighborhood exposure. Linkage was completed through the UCSF Health Atlas, a geospatial platform that integrates UCSF EHR data with nationwide, census tract-level social determinants of health. Marital status was derived from structured EHR fields and collapsed into five categories: Married/Partnered, Single, Divorced/Separated, Widowed, and Unknown/Other. Preferred spoken language (English, Chinese, Spanish, and Other/Non-English) and race/ethnicity (White, Latinx, Black or African American, Asian, Other), were evaluated as a proxy for potential communication or access barriers.(**Figure 1B**).

Cardiovascular risk factors included three continuous exposures: median systolic blood pressure (SBP), median diastolic blood pressure (DBP), and median body mass index (BMI), calculated from all available clinical encounters in the five years prior to or including the date of MCI diagnosis. We also evaluated a dichotomous variable as an indicator of history of type II diabetes at any point in the patient’s EHR prior to MCI diagnosis. These factors were selected based on their established roles in vascular and metabolic risk pathways and their consistent ascertainment within the UCSF EHR (**Figure 1B-C**).

### Covariates

Models evaluating social risk factors (HPI index, marital status, preferred language, race/ethnicity) were adjusted for age at MCI diagnosis, sex, and clinic category. Models evaluating cardiovascular risk factors were adjusted for age at MCI diagnosis, sex, race/ethnicity, social risk factors (preferred spoken language, HPI index, and marital status), and clinic category of the first MCI diagnosis (primary care/internal medicine vs. dementia specialty). When BMI was modeled as the primary exposure, both SBP and DBP were included as covariates to account for cardiovascular burden. We did not adjust for cardiovascular exposures in the models evaluating social risk factors because cardiovascular conditions may be downstream consequences of social determinants of health rather than confounders. Lastly, while all covariates are reported in tables, non-exposure coefficients are not interpreted to avoid the Table 2 fallacy [31].

### Analyses

We used Cox proportional hazards models to estimate the association of social and cardiovascular risk factors with the progression from MCI to AD/ADRD. Two sets of models were constructed: one for social risk factors and one for cardiovascular risk factors (**Figure 1C**). We tested for interactions of sex with both social and cardiovascular risk factors. When an interaction term reached statistical significance (p<0.05), we fit sex-stratified Cox models to estimate hazard ratios separately for men and women.

To assess potential nonlinearity in the relationship between continuous cardiovascular exposures and AD/ADRD progression, we fit restricted cubic spline models for SBP, DBP, and BMI. Linear, quadratic, and cubic spline model fits were compared using the Akaike Information Criterion (AIC) and likelihood ratio tests to inform model fit. Spline models demonstrating a better fit were subsequently visualized with hazard-ratio curves, including sex-stratified splines to explore differences in the shape and magnitude of associations by sex.

## Data Availability

The data used in this study were derived from the UCSF Health electronic health record system and contain protected health information. Due to institutional policies, HIPAA regulations, and the terms of our IRB approval (20-32422), these data are not publicly available. De-identified data may be made available to qualified investigators upon reasonable request, subject to Institutional Review Board approval, completion of data use agreements, and compliance with UCSF data access policies. Statistical analysis code used to generate the results reported below is available from the corresponding author upon reasonable request.

## RESULTS

### Baseline Cohort Characteristics

We included 6,529 patients diagnosed with MCI between 2010 and 2024. Of these, 1,403 (21.5%) progressed to AD/ADRD over the follow-up period. The mean age at MCI diagnosis was 70.9 years (SD=10.0), and 53.5% of the cohort was female. The racial/ethnic composition of the cohort was 55.5% White, 21.1% Asian, 8.0% Latinx, 7.0% Black or African American, and 8.4% identified as another race or ethnicity.

Patients who progressed to dementia were older at baseline (75.2 vs 69.7 years, p <0.001), had higher median SBP (134 mmHg vs. 130 mmHg, p <0.001), lower median DBP (69.5 mmHg vs. 70.8 mmHg, p <0.001), and lower median BMI (25.5 kg/m² vs. 26.1 kg/m², p <0.001) compared to censored individuals. A smaller proportion of those patients who progressed to AD/ADRD had history of type II diabetes diagnosis (63.7% vs. 70.8%, p <0.001). Median follow-up time was substantially shorter among patients who progressed to dementia (281 vs.1,360 days, p<0.001).

Marital status significantly differed (p< 0.001) between groups: 49.0% of those progressing to AD/ADRD were married/partnered compared to 50.6% of censored patients, while widowed individuals comprised 27.8% of individuals who progressed to AD/ADRD compared to 15.3% of those who were censored without an AD/ADRD diagnosis. English was the most common preferred spoken language (83.9% overall) among all patients, followed by Chinese (7.5%), and Spanish (2.5%). The mean California Healthy Places Index (HPI) was similar across AD/ADRD diagnosed patients and those censored (77.2 vs. 77.7), though patients who progressed to AD/ADRD were more likely to have received their MCI diagnosis in a dementia specialty clinic (35.6% vs. 34.1%, p <0.001). Full demographic and clinical characteristics are presented in **Table 1**.

**Table 1.**
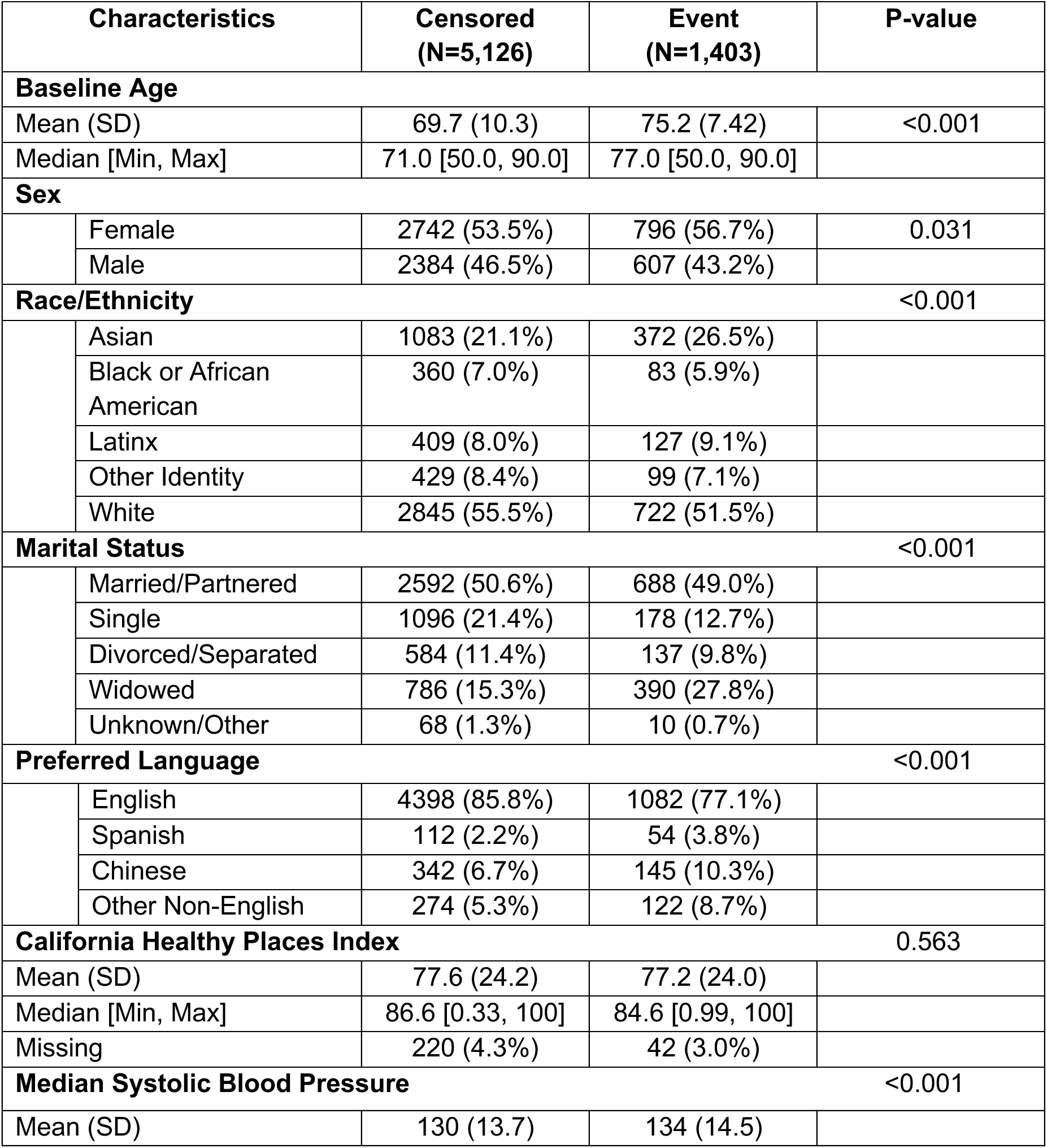
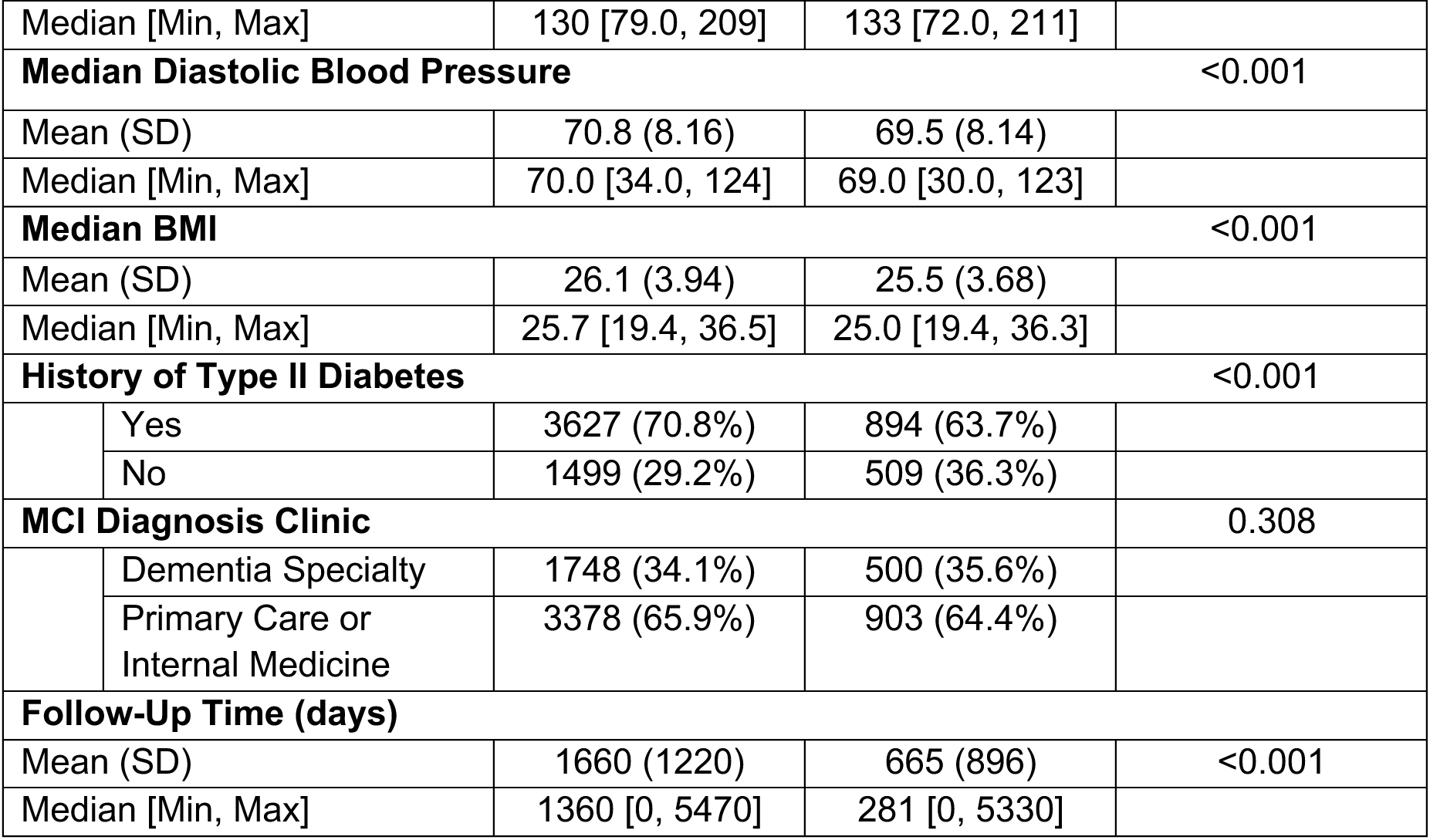
Demographics of Studied Population. Baseline characteristics at the time of MCI diagnosis are presented for individuals who progressed to AD/ADRD (Event) and those who were censored during follow-up. Group comparisons reflect unadjusted differences between Event and Censored participants and were assessed using Welch two-sample t-tests for continuous variables and χ² tests for categorical variables.

| Characteristics |  | Censored<br>(N=5,126) | Event<br>(N=1,403) | P-value |
| --- | --- | --- | --- | --- |
| **Baseline Age** |  |  |  |  |
| Mean (SD) |  | 69.7 (10.3) | 75.2 (7.42) | <0.001 |
| Median [Min, Max] |  | 71.0 [50.0, 90.0] | 77.0 [50.0, 90.0] |  |
| **Sex** |  |  |  |  |
|  | Female | 2742 (53.5%) | 796 (56.7%) | 0.031 |
|  | Male | 2384 (46.5%) | 607 (43.2%) |  |
| **Race/Ethnicity** |  |  |  |  |
|  |  |  |  | <0.001 |
|  | Asian | 1083 (21.1%) | 372 (26.5%) |  |
|  | Black or African American | 360 (7.0%) | 83 (5.9%) |  |
|  | Latinx | 409 (8.0%) | 127 (9.1%) |  |
|  | Other Identity | 429 (8.4%) | 99 (7.1%) |  |
|  | White | 2845 (55.5%) | 722 (51.5%) |  |
| **Marital Status** |  |  |  |  |
|  |  |  |  | <0.001 |
|  | Married/Partnered | 2592 (50.6%) | 688 (49.0%) |  |
|  | Single | 1096 (21.4%) | 178 (12.7%) |  |
|  | Divorced/Separated | 584 (11.4%) | 137 (9.8%) |  |
|  | Widowed | 786 (15.3%) | 390 (27.8%) |  |
|  | Unknown/Other | 68 (1.3%) | 10 (0.7%) |  |
| **Preferred Language** |  |  |  |  |
|  |  |  |  | <0.001 |
|  | English | 4398 (85.8%) | 1082 (77.1%) |  |
|  | Spanish | 112 (2.2%) | 54 (3.8%) |  |
|  | Chinese | 342 (6.7%) | 145 (10.3%) |  |
|  | Other Non-English | 274 (5.3%) | 122 (8.7%) |  |
| **California Healthy Places Index** |  |  |  |  |
|  |  |  |  | 0.563 |
| Mean (SD) |  | 77.6 (24.2) | 77.2 (24.0) |  |
| Median [Min, Max] |  | 86.6 [0.33, 100] | 84.6 [0.99, 100] |  |
| Missing |  | 220 (4.3%) | 42 (3.0%) |  |
| **Median Systolic Blood Pressure** |  |  |  |  |
|  |  |  |  | <0.001 |
| Mean (SD) |  | 130 (13.7) | 134 (14.5) |  |
| Median [Min, Max] |  | 130 [79.0, 209] | 133 [72.0, 211] |  |
| **Median Diastolic Blood Pressure** |  |  |  | <0.001 |
| Mean (SD) |  | 70.8 (8.16) | 69.5 (8.14) |  |
| Median [Min, Max] |  | 70.0 [34.0, 124] | 69.0 [30.0, 123] |  |
| **Median BMI** |  |  |  | <0.001 |
| Mean (SD) |  | 26.1 (3.94) | 25.5 (3.68) |  |
| Median [Min, Max] |  | 25.7 [19.4, 36.5] | 25.0 [19.4, 36.3] |  |
| **History of Type II Diabetes** |  |  |  | <0.001 |
|  | Yes | 3627 (70.8%) | 894 (63.7%) |  |
|  | No | 1499 (29.2%) | 509 (36.3%) |  |
| **MCI Diagnosis Clinic** |  |  |  | 0.308 |
|  | Dementia Specialty | 1748 (34.1%) | 500 (35.6%) |  |
|  | Primary Care or Internal Medicine | 3378 (65.9%) | 903 (64.4%) |  |
| **Follow-Up Time (days)** |  |  |  |  |
| Mean (SD) |  | 1660 (1220) | 665 (896) | <0.001 |
| Median [Min, Max] |  | 1360 [0, 5470] | 281 [0, 5330] |  |

### Social Risk Models Reveal Disparities by Marital Status, Language, and Race/Ethnicity

In fully adjusted Cox proportional hazards models, higher neighborhood advantage was not significantly associated with risk of progression from MCI to AD/ADRD (HR per 10-point increase: 0.99, 95% CI: 0.97–1.01, p=0.33) (**Figure 2)**. Compared to married/partnered individuals, reporting a widowed marital status at baseline was associated with an elevated risk of progression from MCI to AD/ADRD (HR: 1.15; 95% CI: 1.00–1.32; p=0.04); while being single at baseline was associated with a lower hazard of progression (HR: 0.79; 95% CI: 0.67–0.94, p<0.01). No significant associations were observed for patients who were “Divorced/Separated” or were in the “Unknown/Other” marital status group (**Figure 2)**. Age distributions across marital status categories are provided in **eFigure 1** to contextualize potential differences in social and clinical vulnerability.

**Figure 2.**
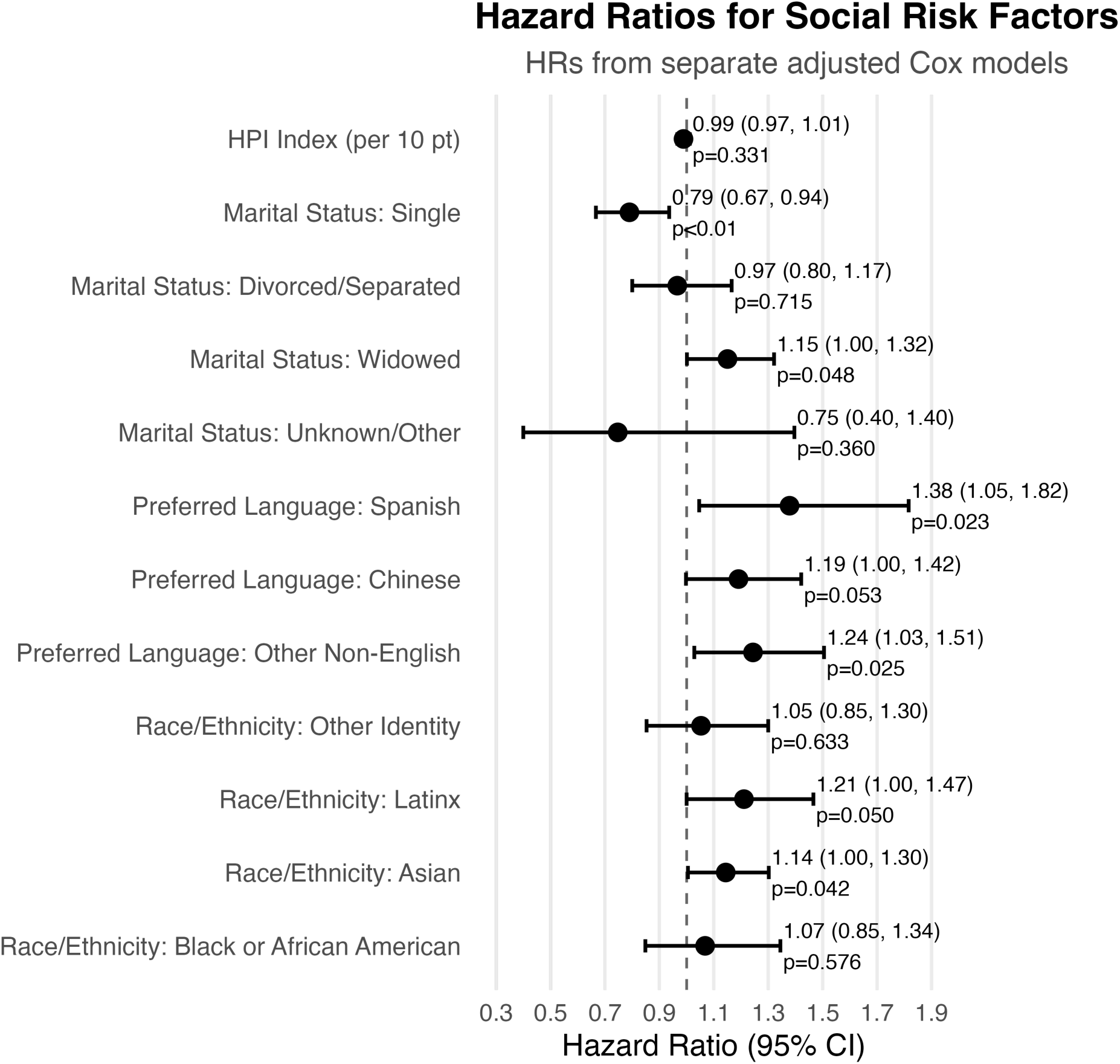
Hazard ratios (HRs) for social risk factors in relation to progression from mild cognitive impairment (MCI) to Alzheimer’s disease and related dementias (ADRD). Each estimate represents the HR and 95% confidence interval from a separate Cox Proportional hazards model, adjusted for age at MCI diagnosis, sex, and other covariates as appropriate. Reference categories are not shown.

Language and race/ethnicity were also associated with risk of progression to dementia. Spanish-speaking patients had a 38% higher hazard of progression compared to English speakers (HR: 1.38; 95% CI: 1.04–1.81, p=0.02). Chinese-speaking patients had a 19% higher hazard of progression compared to English speakers (HR: 1.19; 95% CI: 1.00–1.42; p=0.05), and those with an “Other Non-English” language preference showed a similarly elevated risk (HR: 1.24; 95% CI: 1.03–1.51; p=0.03). In parallel models evaluating race/ethnicity, Latinx (HR:1.21; 95% CI: 1.00–1.47, p=0.05) and Asian patients (HR:1.14, 95% CI: 1.00-1.30; p=0.04) also experienced significantly higher hazards of progression compared to White patients (**Figure 2**).

We found no evidence of sex-based effect modification in the social risk-factors models (**eTables 3-6**).

### SBP is associated with higher and BMI lower hazard of transition to AD/ADRD

In fully adjusted Cox proportional hazards models, higher systolic blood pressure (SBP) was significantly associated with an increased hazard of progression from MCI to AD/ADRD. Each 10 mmHg increase in SBP corresponded to nearly a 10% higher hazard (HR: 1.09, 95% CI: 1.05–1.14; p<0.05). Higher diastolic blood pressure (DBP) was not linearly associated with an elevated hazard of progression. Higher body mass index (BMI) was associated with a lower hazard of progression from MCI to AD/ADRD. Each 1-unit increase in BMI corresponded to a 3% reduced hazard (HR: 0.97; 95% CI: 0.95–0.98; p<0.001). Individuals with a history of type II diabetes averaged slightly faster progression from MCI to AD/ADRD but the confidence interval was too wide to draw conclusions (HR: 1.08, 95% CI: 0.96–1.22; p=0.20) (**Figure 3**).

**Figure 3.**
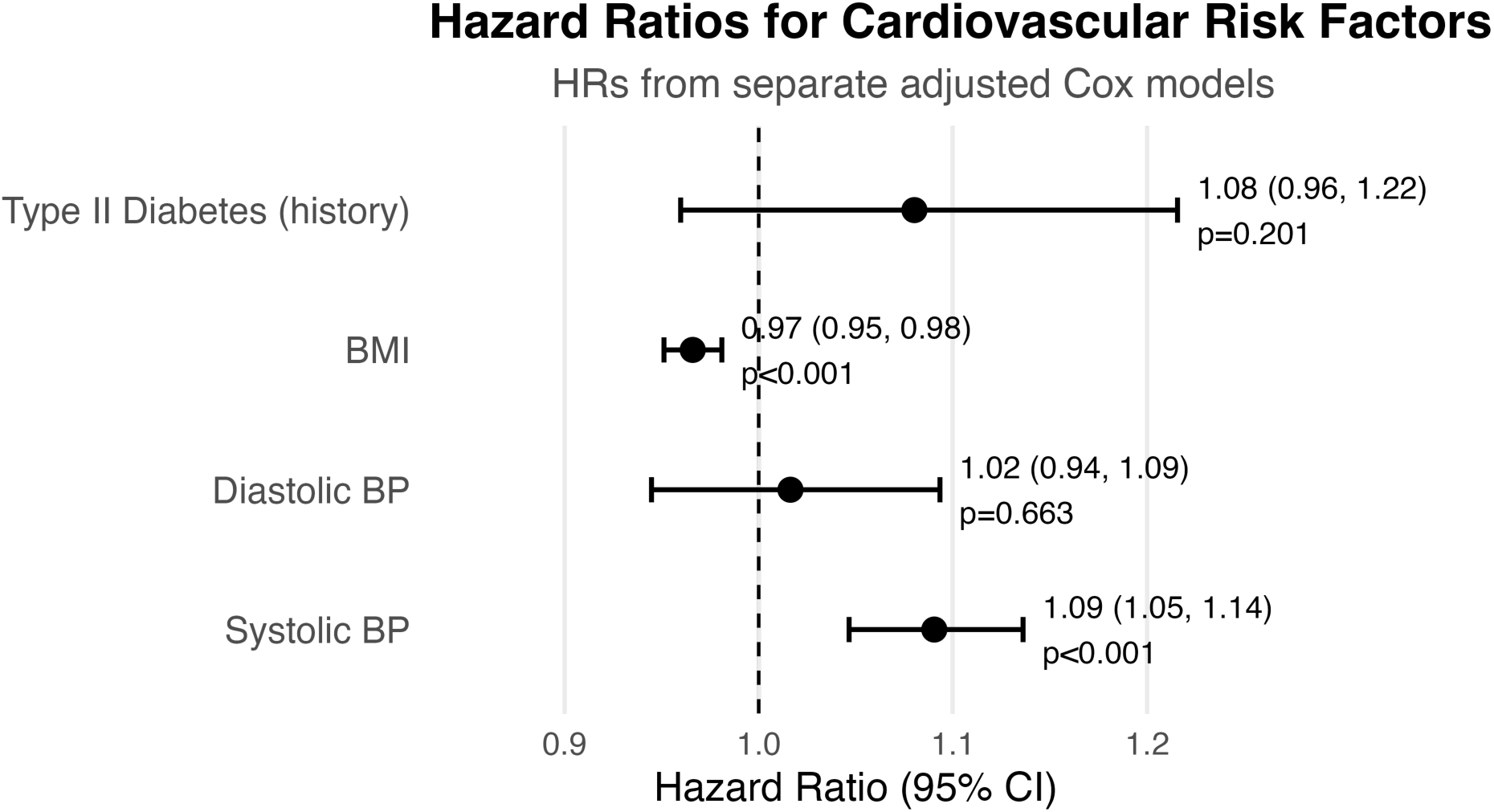
Hazard ratios (HRs) for cardiovascular risk factors and progression from mild cognitive impairment (MCI) to Alzheimer’s disease and related dementias (ADRD). HRs and 95% confidence intervals are shown from separate Cox proportional hazards models for systolic blood pressure (SBP), diastolic blood pressure (DBP), body mass index (BMI), and history of type II diabetes. Each model was adjusted for age at MCI diagnosis, sex, race/ethnicity, and social risk factors (preferred language, race/ethnicity, HPI index, and marital status).

### Sex Interactions Suggest Effect Modification Across All Cardiovascular Risk Factors

The interaction coefficient between SBP and male sex was -0.090 (HR = 0.91; p = 0.03) (**eTables 7-10)**, indicating that the positive association between SBP and dementia progression was attenuated in males compared to females. Among females, each 10 mmHg increase in SBP corresponded to a 13% higher hazard of developing AD/ADRD (HR: 1.13; 95% CI: 1.07–1.19; p <0.001). In contrast, the association was weaker and not statistically significant in males (HR per 10 mmHg: 1.05; 95% CI: 0.98–1.11; p=0.091), showing consistency with the statistically significant interaction between sex and SBP (**Figure 4**).

**Figure 4.**
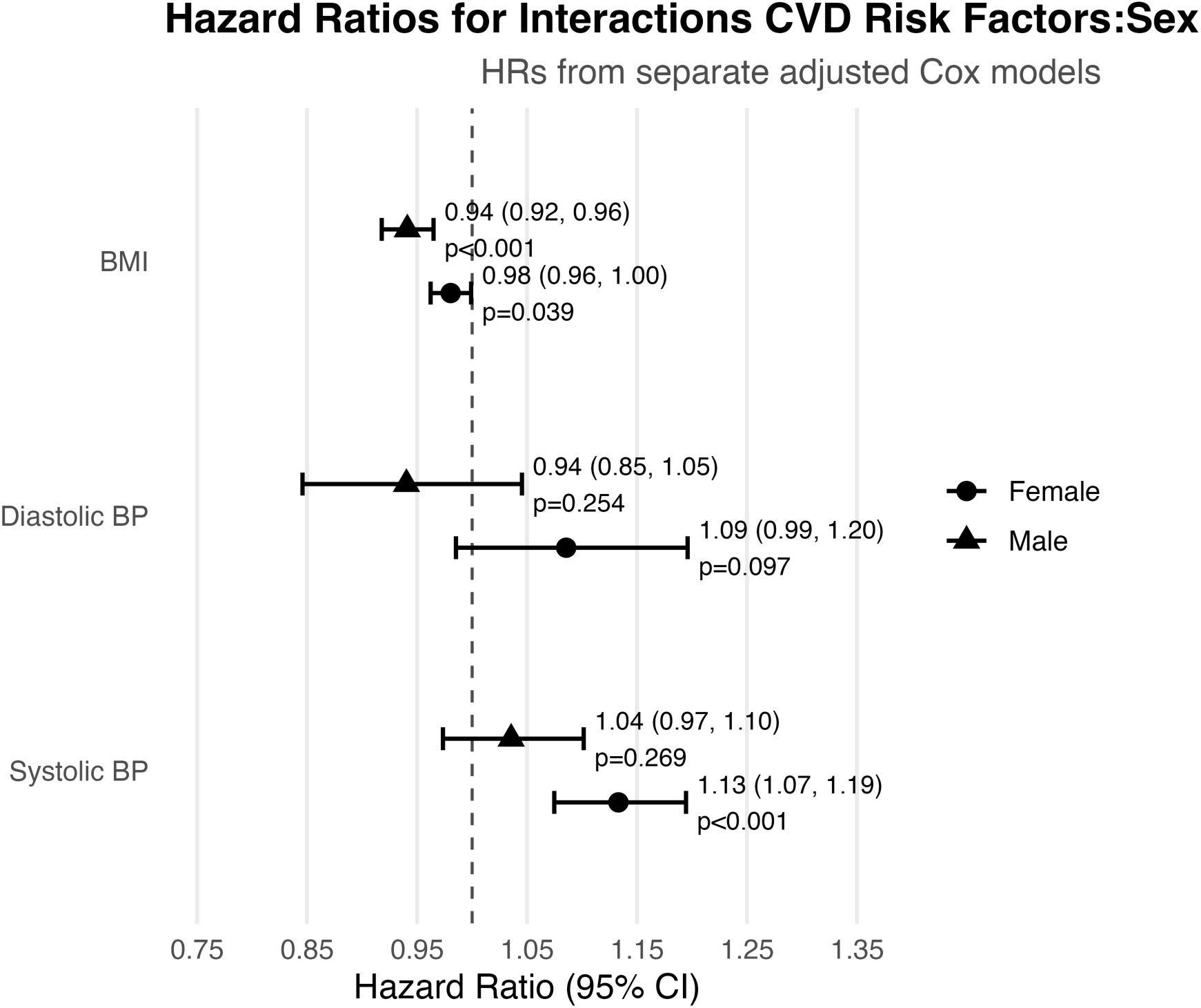
Sex-Stratified hazard ratios for cardiovascular risk factors. Hazard ratios (HRs) and 95% confidence intervals (Cis) from separate Cox proportional hazards models of body mass index (BMI), diastolic blood pressure (DBP), and systolic blood pressure (SBP), stratified by sex. Circles represent estimates for females, triangles for males. Models were adjusted for age at MCI diagnosis, race/ethnicity, marital status, preferred language, HPI index, type II diabetes history, and clinic type.

The interaction term between DBP and male sex was -0.144 (HR = 0.87; p=0.045), suggesting a weaker association in males (**eTable 8**). The sex-stratified models showed non-significant trends in opposite directions. Among females, higher DBP was associated with a non-significant *increase* in risk (HR per 10 mmHg: 1.09; 95% CI: 0.99–1.20; p=0.09), while in males, the association was inverse and not significant (HR per 10 mmHg: 0.93; 95% CI: 0.83–1.04; p=0.214) (**Figure 4**).

The interaction of male sex and BMI (b= -0.041 (HR = 0.96; p = 0.008), indicated that the protective association of higher BMI with reduced AD/ADRD risk was stronger in males than in females (**eTable 9**). Among females, each 1-unit increase in BMI was associated with a 2% reduction in risk (HR:0.98; 95% CI: 0.96–0.99; p=0.022); in males each 1-unit increase in BMI was associated with a 6% reduction in AD/ADRD hazard (HR: 0.94; 95% CI: 0.92–0.97; p<0.001) (**Figure 4**). We found no significant associations between diabetes history and progression risk in either the primary or sex-stratified models, and no evidence of interaction between diabetes and sex (**eTable 10**), reinforcing the overall null finding for diabetes across model specifications.

### Spline Models Improve Model Fit for Blood Pressure but Not BMI

To assess whether flexible spline models improved model fit over linear specifications for each continuous variable, we compared Cox proportional hazards models using both the Akaike Information Criterion (AIC) and Likelihood Ratio Tests (LRT). Lower AIC values indicate better fit, and significant LRT p-values suggest that the spline terms significantly improved model likelihood. Based on these comparisons, spline terms provided best fits for SBP and DBP, while a linear specification was preferred for BMI (**eTable 11**).

### SBP and DBP Show Nonlinear Risk Patterns, BMI Linearly Protective

For SBP, spline models revealed a nonlinear relationship with dementia risk (p<0.05 vs linear model), with increasing hazard beginning at systolic blood pressures above ∼120 mmHg. Risk appeared to plateau between 130-140 mmHg, then rise again in the Stage 2 hypertension range (>140 mmHg) (**Figure 5A** and sex-stratified models in **Figure 6A**). DBP also demonstrated a significant nonlinear association with dementia risk (p<0.05). The hazard ratio remained close to 1.0 across the normal range, with a low point at around 75-80 mmHg. However, risk began to increase steeply beyond 85 mmHg, particularly in the Stage 2 Hypertension range, with wide confidence intervals reflecting increased uncertainty at extreme values (**Figure 5B** and sex-stratified models in **Figure 6B**). In contrast, BMI exhibited a linear inverse association with AD/ADRD risk. Spline modeling did not significantly improve model fit over the linear model specification (p=0.99). The hazard decreased progressively with higher BMI, particularly across the normal to overweight range (20–30 kg/m²) (**eFigure 2,** with sex stratified models in **eFigure 3**).

**Figure 5.**
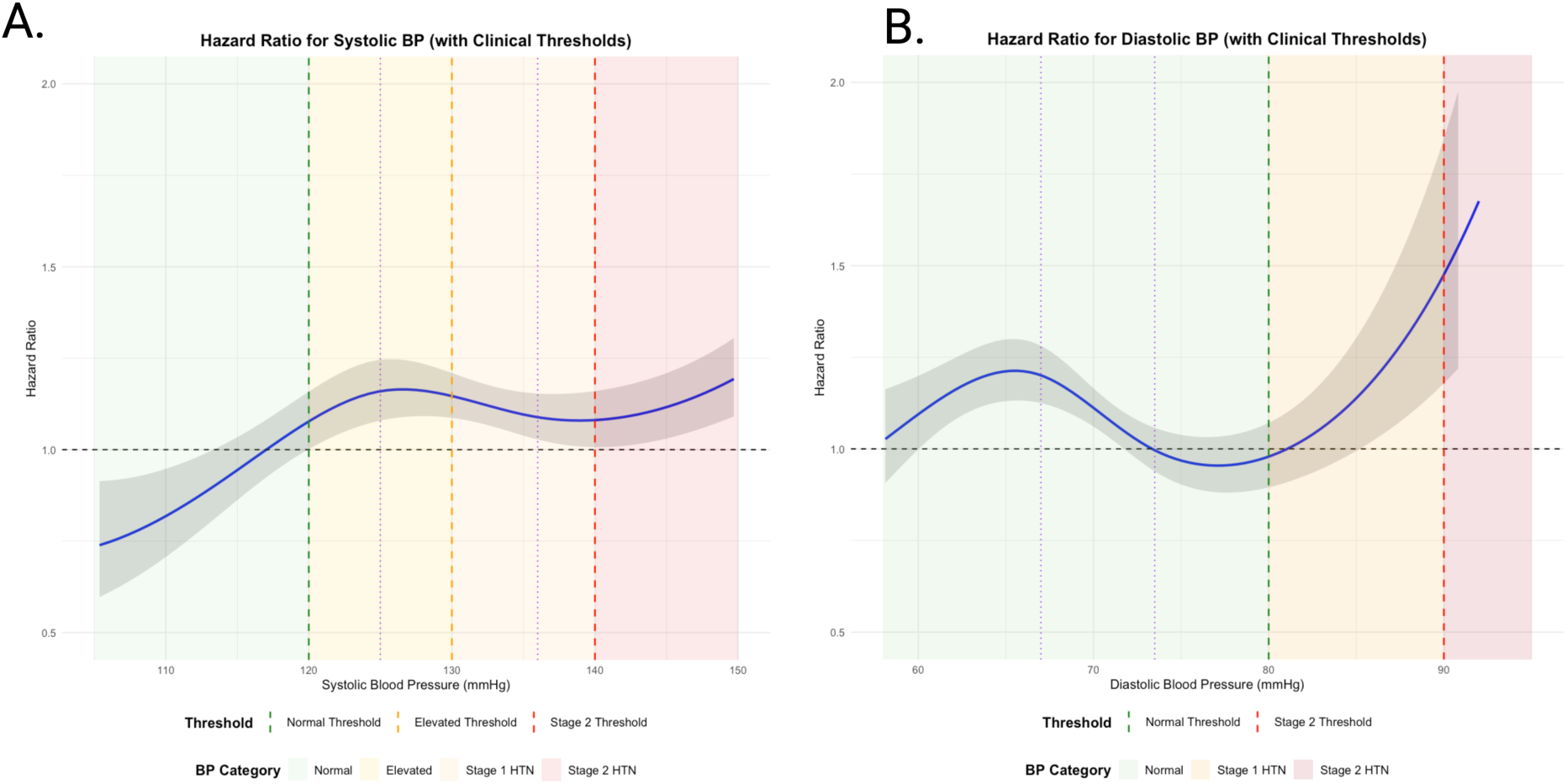
Adjusted hazard ratios for progression to AD/ADRD based on median Systolic (A) and Diastolic (B) Blood Pressure (SBP, DBP), modeled using cubic splines with overlaid clinical thresholds. Shaded regions represent standard U.S. blood pressure categories: normal (green), elevated (yellow), stage 1 hypertension (orange), and stage 2 hypertension (red). Models adjust for age, sex, race/ethnicity, BMI, and social risk factors. Confidence intervals are shaded in gray.

**Figure 6.**
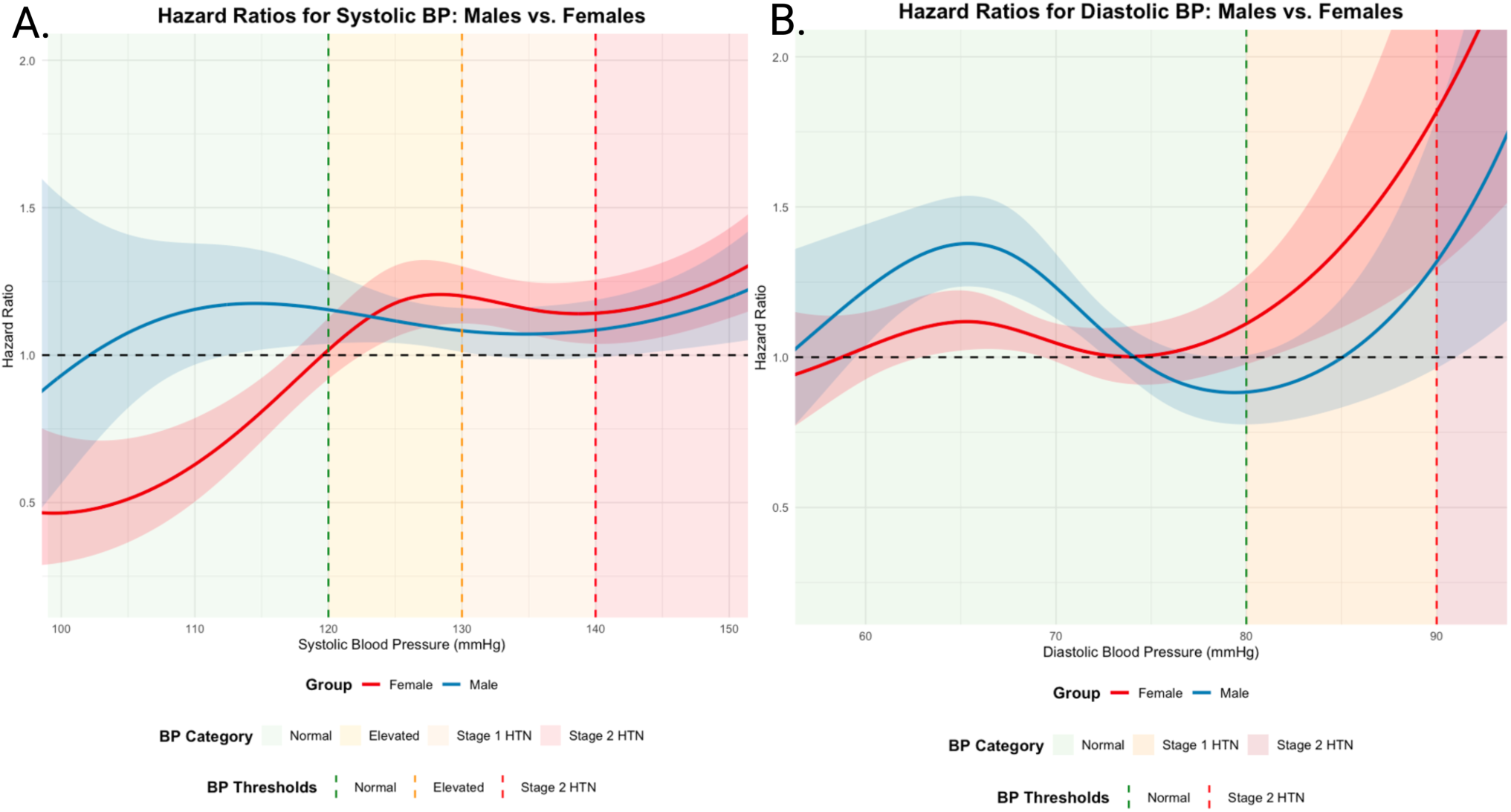
Sex-stratified Spline Models Evaluating the Nonlinear Associations between Systolic and Diastolic Blood Pressure with progression to ADRD. A. Systolic blood pressure (SBP) and B. Diastolic blood pressure (DBP) with risk of progression to Alzheimer’s disease and related dementias (ADRD), adjusted for BMI and Type II diabetes history. Blue lines represent males; red lines represent females. Shaded areas indicate 95% confidence intervals. Colored backgrounds and vertical lines denote clinical blood pressure categories and thresholds per hypertension guidelines: normal (green), elevated (yellow), stage 1 hypertension (orange), and stage 2 hypertension (red).

## DISCUSSION

In this large EHR-based cohort of older adults diagnosed with MCI in real world clinical settings, we found that elevated SBP was significantly associated with progression to AD/ADRD among females, while BMI was associated with lower risk of transition among males. In contrast, DBP showed a nonlinear but less consistent relationship. Social factors such as widowhood, non-English language preference, and minoritized racial/ethnic groups were also associated with increased risk of progression, whereas neighborhood disadvantage was not. Our findings underscore the importance of accessible risk indicators in understanding progression along the AD/ADRD continuum and suggest that structural and social vulnerabilities may influence risk in ways not currently captured by conventional models.

Our results also support prior evidence implicating vascular risk in the rates of progression from MCI to dementia [7,11,32,33] and extend the current literature by demonstrating sex-based differences in both the magnitude and shape of these associations. SBP showed a nonlinear, threshold-like risk pattern beginning at around 120 mmHg, with the steepest rise observed among females, a finding consistent with growing literature on sex-specific vascular contributions to dementia [34–38]. Spline analyses revealed that even SBP values within the prehypertensive range may carry elevated risk for women. These results suggest that standard clinical thresholds may inadequately capture risk among older women with MCI, warranting more nuanced consideration of blood pressure findings in this population.

Although DBP demonstrated nonlinear associations, the effect sizes were smaller and less consistent. Among females, the J-shaped curve hinted at elevated risk at both high and low DBP levels. However, these trends require cautious interpretation and future replication due to wider confidence intervals at the extremes.

BMI exhibited a protective linear association with progression risk, particularly among males. This aligns with population-based studies showing that older individuals with higher late-life BMI average lower risk of dementia. Most interpretations of this finding hypothesize that weight loss is an early indicator of accruing dementia pathology (i.e., undiagnosed or subclinical dementia leads to weight loss), although body weight may also buffer against cognitive decline, possibly via metabolic reserve or frailty pathways [39,40]. While weight gain may not be recommended as a preventative strategy, our findings reinforce the complexity of interpreting BMI in older adults, where lower weight may reflect older age, underlying frailty, or disease burden [41,41–44].

Although prior studies have identified diabetes as a risk factor for dementia incidence and progression from MCI, the estimated hazard ratio in our cohort (HR 1.08) was similar in magnitude to that observed for systolic blood pressure, but with substantially wider confidence intervals. This imprecision limits our ability to draw firm conclusions regarding the role of diabetes in progression risk and suggests that the relationship may depend on cohort characteristics, measurement approach, or downstream effects of clinical management.

While neighborhood disadvantage, measured via the Healthy Places Index (HPI), was not associated with risk of progression in our models, other social exposures, namely preferred language and being widowed, were significant predictors. Patients who preferred a non-English language had 19–38% higher hazards of progression, independent of age, sex, and clinic type. These associations may reflect barriers to accessing or navigating post-MCI care, as well as potential underdiagnosis or delayed detection of MCI due to linguistic mismatch between providers and patients. The lack of association of HPI with progression to dementia may reflect the relatively high average neighborhood advantage in our cohort or limitations in census tracts in capturing individual-level adversity. Alternatively, structural disadvantage may shape earlier stages of cognitive vulnerability (e.g., lower education attainment), while clinical progression after MCI may be driven more by individual health management.

Widowed patients also faced elevated risk, consistent with literature highlighting the role of spousal support in cognitive health [45]. Notably, the association did not vary by sex, suggesting that widowhood may reduce social support or increase psychological stress regardless of gender. In contrast, patients who were single had a lower risk of progression, indicating that the observed association was not attributable to being unmarried per se. One interpretation is that individuals who have never married may have established alternative social support structures over time, whereas widowhood may represent a more abrupt disruption to existing support systems and psychological wellbeing. However, we lacked information on the timing of widowhood, which limits our ability to assess how recency of spousal loss may influence progression risk.

Both Latinx and Asian patients exhibited significantly higher hazards of progression compared to White patients, after adjusting for age, sex, and clinical setting. These disparities are concerning and may reflect differences in comorbidity management, delays in MCI diagnoses, or exposure to chronic stressors not captured in their EHR data. Since disparities persist despite adjustment for preferred language, this suggests that sociocultural and structural factors beyond linguistic access may contribute to inequities and disease progression.

Our findings add to a small but growing body of research examining racial and ethnic disparities in post-MCI outcomes. Future work is needed to disentangle the relative contributions of biological, behavioral, and structural drivers of these inequities, particularly given that many predictive models fail to include social determinants or culturally responsive measures. A key strength of our study is its emphasis on routinely collected EHR variables: blood pressure, BMI, diabetes history, preferred spoken language, and marital status, which are often overlooked in prediction models, and instead rely on clinically meaningful advanced imaging and biomarkers [46]. These findings support the development of more accessible, equitable risk stratification tools that can be deployed across diverse clinical settings when high-resource diagnostics are not available. Our sex-stratified and spline-based models further demonstrate the value of flexible approaches to clinical risk estimation, which may enhance precision in MCI care planning.

Our study has several limitations. First, the observational nature of EHR data limits our ability to make causal inferences, and residual confounding is possible despite our covariate adjustment. Second, our exposure measures, such as blood pressure and BMI, were calculated as median values over a five-year window, based on available measurements in the EHR. This approach was necessary given that structured UCSF EHR data were only consistently available beginning in 2010, but it may not fully capture acute fluctuations, long-term variability, or the timing of exposure relative to diagnosis. Third, we did not account for clinical treatment or medication management of key exposures, such as antihypertensive use or glycemic control, which may influence both the cardiovascular exposures and dementia risk. Fourth, our analyses treated cardiovascular and social variables as baseline exposures, although these characteristics may change over time. It is possible that new-onset conditions (e.g., incident diabetes) or changing social circumstances (e.g., widowhood) during the MCI-to-AD/ADRD transition may influence risk in ways not captured in this study. Fifth, while we attempted to incorporate structural factors using neighborhood level HPI-scores, our cohort was largely drawn from the San Francisco Bay Area, where most patients reside in relatively advantaged neighborhoods. This limited variability may have reduced our ability to detect associations between neighborhood disadvantage and dementia progression. Sixth, we did not address missing data directly in our modeling approach, and exposure ascertainment depended on the availability of sufficient clinical data for each patient. Finally, MCI and dementia diagnoses were determined based on ICD codes documented in the EHR, which are primarily used for billing and administrative purposes, limiting insights into underlying etiology. As such, they may not accurately reflect cognitive status due to limited availability of cognitive test results, systematic measurement of functional decline, or biomarker confirmation in EHR data.

In sum, our study highlights the importance of both clinical and social risk factors in shaping the transition from MCI to AD/ADRD. Elevated SBP, low BMI, widowhood, linguistic barriers, and racial/ethnic minority status all emerged as significant predictors of progression. Notably, the protective association of higher BMI was stronger among men, while elevated SBP was more strongly linked to risk in women. These findings point to the need for equitable approaches to dementia risk stratification and the utility of leveraging real-world data to understand prognosis among individuals living with MCI.

