## Supplemental Materials for "Social and Cardiovascular Risk Factors as Predictors of the Progression from Mild Cognitive Impairment to Dementia in a Large EHR Database"

**Supplemental eTable T1. ICD Codes for MCI Diagnoses.** ICD Codes include ICD-9 and ICD-10 codes used for MCI diagnoses with included description.

| ICD Code | Description |
|---|---|
| G31.84 | Mild cognitive impairment of uncertain or unknown etiology |
| R41.1 | Anterograde amnesia |
| R41.2 | Retrograde amnesia |
| R41.3 | Other amnesia |
| R41.81 | Age-related cognitive decline |
| R41.82 | Altered mental status, unspecified |
| R41.840 | Attention and concentration deficit |
| R41.841 | Cognitive communication deficit |
| R41.842 | Visuospatial deficit |
| R41.843 | Psychomotor deficit |
| R41.844 | Frontal lobe and executive function deficit |
| R41.89 | Other symptoms and signs involving cognitive functions and awareness |
| R41.9 | Unspecified symptoms and signs involving cognitive functions and awareness |
| F06.8 | Other specified mental disorders due to known physiological condition |
| 331.83 | Mild Cognitive Impairment of uncertain or unknown etiology |
| 780.93 | Memory Loss |
| 797 | Senility without mention of psychosis |
| 780.97 | Altered mental status |
| 799.51 | Attention and concentration deficit |
| 799.52 | Unspecified signs and symptoms involving cognition |
| 799.53 | Visuospatial deficit |
| 799.54 | Psychomotor deficit |
| 799.55 | Frontal lobe and executive function deficit |
| 799.59 | Other signs and symptoms involving cognition |
| 294.8 | Other persistent mental disorders due to conditions classified elsewhere |

**Supplemental eTable T2. ICD Codes for AD/ADRD Diagnoses**
ICD Codes include ICD-9 and ICD-10 codes used for AD/ADRD diagnoses with included description.

| ICD Code | Description |
|---|---|
| F00.0 | Dementia in Alzheimer disease |
| F00.1 | Dementia in Alzheimer disease with late onset |
| F00.2 | Dementia in Alzheimer's disease, atypical or mixed type |
| F00.9 | Dementia in Alzheimer's disease, unspecified |
| F01 | Vascular Dementia |
| F01.5 | Vascular Dementia, unspecified severity |
| F01.50 | Vascular Dementia with behavioral disturbance, psychotic disturbance, mood disturbance, and anxiety |
| F01.51 | Vascular Dementia, unspecified severity, with unspecified severity, with agitation |
| F01.518 | Vascular dementia, unspecified severity, with other behavioral disturbance. |
| F01.52 | Vascular Dementia with psychotic disturbance |
| F01.53 | Vascular Dementia with mood disturbance |
| F01.54 | Vascular Dementia with Anxiety |
| F01.A | Vascular Dementia, mild |
| F01.A0 | Vascular Dementia mild, without behavioral disturbance, psychotic disturbance, mood disturbance and anxiety |
| F01.A1 | Vascular Dementia, mild, with behavioral disturbance |
| F01.A11 | Vascular Dementia, mild, with agitation |
| F01.A18 | Vascular dementia, mild, with other behavioral disturbance |
| F01.A2 | Vascular dementia with psychotic disturbance |
| F01.A3 | Vascular dementia with mood disturbance |
| F01.A4 | Vascular Dementia with anxiety |
| F01.B | Vascular Dementia, moderate |
| F01.B0 | Vascular Dementia, moderate, without behavioral disturbance, psychotic disturbance, mood disturbance and anxiety |
| F01.B1 | Vascular Dementia, moderate, with behavioral disturbance |
| F01.B11 | Vascular Dementia, moderate, with agitation |
| F01.B18 | Vascular Dementia, moderate, with other behavioral disturbance |
| F01.B2 | Vascular dementia, moderate, with psychotic disturbance |
| F01.B3 | Vascular Dementia, moderate, with mood disturbance |
| F01.B4 | Vascular Dementia, moderate, with anxiety |
| F01.C | Vascular Dementia, severe |
| F01.C0 | Vascular Dementia, severe, without behavioral disturbance, psychotic disturbance, mood disturbance, and anxiety |
| F01.C1 | Vascular Dementia, severe, with behavioral disturbance |

| ICD Code | Description |
|---|---|
| F01.C11 | Vascular Dementia, severe, with agitation |
| F01.C18 | Vascular Dementia, severe, with other behavioral disturbance |
| F01.C2 | Vascular Dementia, severe, with psychotic disturbance |
| F01.C3 | Vascular Dementia, severe, with mood disturbance |
| F01.C4 | Vascular Dementia, severe, with anxiety |
| F01.2 | Subcortical Vascular Dementia |
| F01.3 | Mixed Cortical and subcortical vascular dementia |
| F01.8 | Other Vascular Dementia |
| F02 | Dementia in other diseases classified elsewhere |
| F02.8 | Dementia in other diseases classified elsewhere, unspecified severity |
| F02.80 | Dementia in other diseases classified elsewhere, without behavioral disturbance, psychotic disturbance, mood disturbance, and anxiety |
| F02.81 | Dementia in other diseases classified elsewhere, unspecified severity with behavioral disturbance |
| F02.811 | Dementia in other disease classified elsewhere, unspecified severity, with agitation |
| F02.818 | Dementia in other disease classified elsewhere, unspecified severity, with other behavioral disturbance |
| F02.82 | Dementia in other diseases classified elsewhere with psychotic disturbance |
| F02.83 | Dementia in other diseases classified elsewhere with mood disturbance |
| F02.84 | Dementia in other diseases classified elsewhere with anxiety |
| F02.A | Dementia in other diseases classified elsewhere, mild |
| F02.A0 | Dementia in other diseases classified elsewhere, mild, without behavioral disturbance, psychotic disturbance, mood disturbance, and anxiety |
| F02.A1 | Dementia in other diseases classified elsewhere, mild, with behavioral disturbance |
| F02.A11 | Dementia in other diseases classified elsewhere, mild, with agitation |
| F02.A18 | Dementia in other diseases classified elsewhere, mild, with other behavioral disturbance |
| F02.A2 | Dementia in other diseases classified elsewhere, mild, with psychotic disturbance |
| F02.A3 | Dementia in other diseases classified elsewhere, mild, with mood disturbance |
| F02.A4 | Dementia in other diseases classified elsewhere, mild, with anxiety |
| F02.B | Dementia in other diseases classified elsewhere, moderate |

| ICD Code | Description |
| --- | --- |
| F02.B0 | Dementia in other diseases classified elsewhere, moderate, without behavioral disturbance, mood disturbance, and anxiety |
| F02.B1 | Dementia in other diseases classified elsewhere, moderate, with behavioral disturbance |
| F02.B11 | Dementia in other diseases classified elsewhere, moderate, with agitation |
| F02.B18 | Dementia in other diseases classified elsewhere, moderate, with other behavioral disturbance |
| F02.B2 | Dementia in other diseases classified elsewhere, moderate, with psychotic disturbance |
| F02.B3 | Dementia in other diseases classified elsewhere, moderate, with mood disturbance |
| F02.B4 | Dementia in other diseases classified elsewhere, moderate, with anxiety |
| F02.C | Dementia in other diseases classified elsewhere, severe |
| F02.C0 | Dementia in other diseases classified elsewhere, severe, without behavioral disturbance, psychotic disturbance, mood disturbance, and anxiety |
| F02.C1 | Dementia in other diseases classified elsewhere, severe, with behavioral disturbance |
| F02.C11 | Dementia in other diseases classified elsewhere, severe, with agitation |
| F02.C18 | Dementia in other diseases classified elsewhere, severe, with other behavioral disturbance. |
| F02.C2 | Dementia in other diseases classified elsewhere, severe, with psychotic disturbance |
| F02.C3 | Dementia in other diseases classified elsewhere, severe, with mood disturbance |
| F02.C4 | Dementia in other diseases classified elsewhere, severe, with anxiety |
| F03 | Unspecified Dementia |
| F03.9 | Unspecified dementia, unspecified severity |
| F03.90 | Unspecified dementia, unspecified severity without behavioral disturbance, psychotic disturbance, mood disturbance, and anxiety |
| F03.91 | Unspecified dementia, unspecified severity, with behavioral disturbance |
| F03.911 | Unspecified dementia, unspecified severity, with agitation |
| F03.918 | Unspecified dementia, unspecified severity, with other behavioral disturbance. |
| F03.92 | Unspecified dementia, unspecified severity with psychotic disturbance |
| F03.93 | Unspecified dementia, unspecified severity with mood disturbance |
| F03.94 | Unspecified dementia, unspecified severity with anxiety |

| ICD Code | Description |
|---|---|
| F03.A | Unspecified dementia, mild |
| F03.A0 | Unspecified dementia, mild without behavioral disturbance, psychotic disturbance, mood disturbance, and anxiety. |
| F03.A1 | Unspecified dementia, mild, with behavioral disturbance |
| F03.A11 | Unspecified dementia, mild, with agitation |
| F03.A18 | Unspecified dementia, mild, with other behavioral disturbance |
| F03.A2 | Unspecified dementia, mild, with psychotic disturbance |
| F03.A3 | Unspecified dementia, mild, with mood disturbance |
| F03.A4 | Unspecified dementia, mild, with anxiety |
| F03.B | Unspecified dementia, moderate |
| F03.B0 | Unspecified dementia, moderate, without behavioral disturbance, psychotic disturbance, mood disturbance, and anxiety |
| F03.B1 | Unspecified dementia, moderate, with behavioral disturbance |
| F03.B11 | Unspecified dementia, moderate, with agitation |
| F03.B18 | Unspecified dementia, moderate, with other behavioral disturbance |
| F03.B2 | Unspecified dementia, moderate, with psychotic disturbance |
| F03.B3 | Unspecified dementia, moderate, with mood disturbance |
| F03.B4 | Unspecified dementia, moderate, with anxiety |
| F03.C | Unspecified dementia, severe |
| F03.C0 | Unspecified dementia, severe, without behavioral disturbance, psychotic disturbance, mood disturbance, and anxiety |
| F03.C1 | Unspecified dementia, severe, with behavioral disturbance |
| F03.C11 | Unspecified dementia, severe, with agitation |
| F03.C18 | Unspecified dementia, severe, with other behavioral disturbance |
| F03.C2 | Unspecified dementia, severe, with psychotic disturbance |
| F03.C3 | Unspecified dementia, severe, with mood disturbance |
| F03.C4 | Unspecified dementia, severe, with anxeity |
| G30 | Alzheimer's Disease |
| G30.0 | Alzheimer's Disease with early onset |
| G30.1 | Alzheimer's Disease with late onset |
| G30.8 | Other Alzheimer's Disease |
| G30.9 | Alzheimer's Disease, unspecified |
| G31.0 | Frontotemporal Dementia |
| G31.01 | Pick's Disease |
| G31.1 | Senile degeneration of brain, not elsewhere classified |
| G31.09 | Other Frontotemporal neurocognitive disorder |
| G31.83 | Neurocognitive Disorder with Lewy bodies |

| **ICD Code** | **Description** |
| --- | --- |
| 290 . 0 | Senile dementia, simple type |
| 290.1 | Presenile dementia |
| 290.2 | Senile dementia, depressed or paranoid type |
| 290.3 | Unspecified dementia, unspecified severity, without behavioral disturbance, psychotic disturbance, mood disturbance, and anxiety |
| 290.4 | Vascular dementia, unspecified severity, without behavioral disturbance |
| 290.4 0 | Vascular dementia, uncomplicated |
| 290.41 | Vascular dementia, unspecified severity, with behavioral disturbance |
| 290.9 | Unspecified dementia, unspecified severity, without behavioral disturbance, psychotic disturbance, mood disturbance, and anxiety |
| 294.1 0 | Dementia in other diseases classified elsewhere, unspecified severity, without behavioral disturbance, psychotic disturbance, mood disturbance, and anxiety |
| 294.11 | Dementia in conditions classified elsewhere with behavioral disturbance |
| 294.2 | Dementia with no underlying cause or specified cause. |
| 294.2 0 | Unspecified dementia, unspecified severity, without behavioral disturbance, psychotic disturbance, mood disturbance, and anxiety |
| 294.21 | Dementia with unspecified severity and behavioral disturbance |
| 331. 0 | Alzheimer's Disease |
| 331.2 | Senile degeneration of the brain |
| 331.8 | Cerebral Degeneration |
| 331.19 | Other frontotemporal neurocognitive disorder |
| 331.82 | Neurocognitive disorder with Lewy bodies |

**Supplemental eTable T3. HPI × Sex Interaction**

| Term | Beta | Hazard Ratio | 95% CI | p-value |
|---|---|---|---|---|
| HPI Index (per 10 pt) | -0.013 | 0.99 | (0.96, 1.02) | 0.374 |
| **HPI Index × Male** | **-0.004** | **1.00** | **(0.95, 1.04)** | **0.877** |
| Sex: Male | -0.004 | 1.00 | (0.69, 1.44) | 0.981 |
| Age at MCI | 0.063 | 1.07 | (1.06, 1.07) | <0.001 |
| Clinic Category: Primary Care or Internal Medicine | -0.181 | 0.83 | (0.74, 0.94) | 0.003 |
| Marital Status: Divorced/Separated | -0.004 | 1.00 | (0.82, 1.21) | 0.970 |
| Marital Status: Single | -0.249 | 0.78 | (0.66, 0.93) | 0.005 |
| Marital Status: Unknown/Other | -0.441 | 0.64 | (0.32, 1.29) | 0.216 |
| Marital Status: Widowed | 0.145 | 1.16 | (1, 1.33) | 0.043 |
| Preferred Language: Chinese | 0.100 | 1.10 | (0.89, 1.37) | 0.367 |
| Preferred Language: Other Non-English | 0.147 | 1.16 | (0.94, 1.42) | 0.163 |
| Preferred Language: Spanish | 0.144 | 1.15 | (0.81, 1.64) | 0.423 |
| Race/Ethnicity: Asian | 0.038 | 1.04 | (0.89, 1.22) | 0.640 |
| Race/Ethnicity: Black or African American | 0.059 | 1.06 | (0.84, 1.35) | 0.630 |
| Race/Ethnicity: Latinx | 0.119 | 1.13 | (0.89, 1.43) | 0.324 |
| Race/Ethnicity: Other Identity | 0.025 | 1.03 | (0.83, 1.27) | 0.818 |

**Supplemental eTable T4. Marital Status × Sex Interaction**

| Term | Beta | Hazard Ratio | 95% CI | p-value |
|---|---|---|---|---|
| **Marital Status (Divorced/Separated) × Male** | **0.005** | **1.01** | **(0.67, 1.5)** | **0.979** |
| **Marital Status (Single) × Male** | **-0.254** | **0.78** | **(0.55, 1.09)** | **0.143** |
| **Marital Status (Unknown/Other) × Male** | **0.379** | **1.46** | **(0.42, 5.1)** | **0.552** |
| **Marital Status (Widowed) × Male** | **0.049** | **1.05** | **(0.78, 1.41)** | **0.743** |
| Marital Status: Divorced/Separated | -0.029 | 0.97 | (0.77, 1.23) | 0.805 |
| Marital Status: Single | -0.129 | 0.88 | (0.7, 1.1) | 0.258 |
| Marital Status: Unknown/Other | -0.459 | 0.63 | (0.26, 1.53) | 0.309 |
| Marital Status: Widowed | 0.141 | 1.15 | (0.97, 1.36) | 0.101 |
| Sex: Male | -0.019 | 0.98 | (0.84, 1.14) | 0.809 |
| Age at MCI | 0.063 | 1.06 | (1.06, 1.07) | <0.001 |
| Clinic Category: Primary Care or Internal Medicine | -0.200 | 0.82 | (0.73, 0.92) | <0.001 |
| Preferred Language: Chinese | 0.128 | 1.14 | (0.92, 1.41) | 0.242 |
| Preferred Language: Other Non-English | 0.182 | 1.20 | (0.98, 1.47) | 0.078 |
| Preferred Language: Spanish | 0.247 | 1.28 | (0.91, 1.8) | 0.156 |
| Race/Ethnicity: Asian | 0.035 | 1.04 | (0.88, 1.21) | 0.669 |
| Race/Ethnicity: Black or African American | 0.090 | 1.09 | (0.87, 1.38) | 0.448 |
| Race/Ethnicity: Latinx | 0.104 | 1.11 | (0.88, 1.4) | 0.383 |
| Race/Ethnicity: Other Identity | 0.011 | 1.01 | (0.82, 1.25) | 0.919 |

**Supplemental eTable T5. Preferred Language × Sex Interaction**

| Term | Beta | Hazard Ratio | 95% CI | p-value |
|---|---|---|---|---|
| **Language (Chinese) × Male** | **-0.131** | **0.88** | **(0.61, 1.26)** | **0.475** |
| **Language (Other Non-English) × Male** | **-0.277** | **0.76** | **(0.5, 1.15)** | **0.197** |
| **Language (Spanish) × Male** | **-0.417** | **0.66** | **(0.34, 1.27)** | **0.211** |
| Preferred Language: Chinese | 0.226 | 1.25 | (1.01, 1.56) | 0.045 |
| Preferred Language: Other Non-English | 0.305 | 1.36 | (1.08, 1.7) | 0.008 |
| Preferred Language: Spanish | 0.436 | 1.55 | (1.13, 2.12) | 0.007 |
| Sex: Male | -0.032 | 0.97 | (0.86, 1.09) | 0.599 |
| Age at MCI | 0.067 | 1.07 | (1.06, 1.08) | <0.001 |
| Clinic Category: Primary Care or Internal Medicine | -0.189 | 0.83 | (0.74, 0.93) | <0.001 |

**Supplemental eTable T6. Race/Ethnicity × Sex Interaction**

| Term | Beta | Hazard Ratio | 95% CI | p-value |
|---|---|---|---|---|
| **Race/Ethnicity (Asian) × Male** | **-0.011** | **0.99** | **(0.77, 1.28)** | **0.935** |
| **Race/Ethnicity (Black or African American) × Male** | **-0.390** | **0.68** | **(0.41, 1.13)** | **0.133** |
| **Race/Ethnicity (Latinx) × Male** | **-0.178** | **0.84** | **(0.55, 1.27)** | **0.399** |
| **Race/Ethnicity (Other Identity) × Male** | **-0.059** | **0.94** | **(0.61, 1.44)** | **0.786** |
| Race/Ethnicity: Asian | 0.143 | 1.15 | (0.97, 1.36) | 0.097 |
| Race/Ethnicity: Black or African American | 0.190 | 1.21 | (0.92, 1.59) | 0.173 |
| Race/Ethnicity: Latinx | 0.252 | 1.29 | (1.02, 1.62) | 0.033 |
| Race/Ethnicity: Other Identity | 0.079 | 1.08 | (0.82, 1.43) | 0.576 |
| Sex: Male | -0.037 | 0.96 | (0.83, 1.12) | 0.617 |
| Age at MCI | 0.068 | 1.07 | (1.06, 1.08) | <0.001 |
| Clinic Category: Primary Care or Internal Medicine | -0.186 | 0.83 | (0.74, 0.93) | 0.001 |

**Supplemental eTable T7. Sex Interaction Coefficient with Systolic Blood Pressure.**
The final row reports the interaction coefficient, while preceding rows show covariate-adjusted hazard ratios for progression to dementia.

| Term | β (coef) | SE | Hazard Ratio | 95% CI | p-value |
|---|---|---|---|---|---|
| Systolic BP | 0.125 | 0.027 | 1.13 | (1.07, 1.19) | <0.001 |
| Sex: Male | 1.194 | 0.543 | 3.30 | (1.14, 9.57) | 0.028 |
| BMI | -0.035 | 0.008 | 0.97 | (0.95, 0.98) | <0.001 |
| Type II Diabetes (history) | 0.092 | 0.060 | 1.10 | (0.97, 1.23) | 0.125 |
| Age at MCI | 0.057 | 0.004 | 1.06 | (1.05, 1.07) | <0.001 |
| Race/Ethnicity: Other Identity | 0.010 | 0.111 | 1.01 | (0.81, 1.26) | 0.929 |
| Race/Ethnicity: Latinx | 0.153 | 0.121 | 1.17 | (0.92, 1.48) | 0.206 |
| Race/Ethnicity: Asian | -0.027 | 0.083 | 0.97 | (0.83, 1.15) | 0.748 |
| Race/Ethnicity: Black or African American | 0.055 | 0.123 | 1.06 | (0.83, 1.35) | 0.654 |
| Marital Status: Married/Partnered | 0.013 | 0.097 | 1.01 | (0.84, 1.23) | 0.891 |
| Marital Status: Single | -0.249 | 0.116 | 0.78 | (0.62, 0.98) | 0.032 |
| Marital Status: Unknown/Other | -0.436 | 0.365 | 0.65 | (0.32, 1.32) | 0.232 |
| Marital Status: Widowed | 0.138 | 0.103 | 1.15 | (0.94, 1.41) | 0.182 |
| Preferred Language: Spanish | 0.117 | 0.179 | 1.12 | (0.79, 1.60) | 0.512 |
| Preferred Language: Chinese | 0.105 | 0.110 | 1.11 | (0.89, 1.38) | 0.344 |
| Preferred Language: Other Non-English | 0.166 | 0.106 | 1.18 | (0.96, 1.45) | 0.115 |
| Clinic Category: Primary Care or Internal Medicine | -0.183 | 0.060 | 0.83 | (0.74, 0.94) | 0.002 |
| HPI Index (per 10 pt) | -0.014 | 0.012 | 0.99 | (0.96, 1.01) | 0.254 |
| **Systolic BP × Male** | **-0.090** | **0.041** | **0.91** | **(0.84, 0.99)** | **0.027** |

**Supplemental eTable T8. Sex Interaction Coefficient with Diastolic Blood Pressure.** The final row reports the interaction coefficient, while preceding rows show covariate-adjusted hazard ratios for progression to dementia.

| Term | β (coef) | SE | Hazard Ratio | 95% CI | p-value |
|---|---|---|---|---|---|
| Diastolic BP | 0.082 | 0.049 | 1.09 | (0.99, 1.20) | 0.097 |
| Sex: Male | 0.988 | 0.504 | 2.69 | (1.00, 7.21) | 0.050 |
| BMI | -0.031 | 0.008 | 0.97 | (0.95, 0.98) | <0.001 |
| Type II Diabetes (history) | 0.097 | 0.060 | 1.10 | (0.98, 1.24) | 0.109 |
| Age at MCI | 0.061 | 0.004 | 1.06 | (1.06, 1.07) | <0.001 |
| Race/Ethnicity: Other Identity | 0.024 | 0.111 | 1.02 | (0.82, 1.27) | 0.829 |
| Race/Ethnicity: Latinx | 0.146 | 0.122 | 1.16 | (0.91, 1.47) | 0.230 |
| Race/Ethnicity: Asian | -0.021 | 0.083 | 0.98 | (0.83, 1.15) | 0.805 |
| Race/Ethnicity: Black or African American | 0.085 | 0.123 | 1.09 | (0.86, 1.38) | 0.492 |
| Marital Status: Married/Partnered | 0.008 | 0.097 | 1.01 | (0.83, 1.22) | 0.932 |
| Marital Status: Single | -0.241 | 0.116 | 0.79 | (0.63, 0.99) | 0.038 |
| Marital Status: Unknown/Other | -0.425 | 0.365 | 0.65 | (0.32, 1.34) | 0.244 |
| Marital Status: Widowed | 0.163 | 0.104 | 1.18 | (0.96, 1.44) | 0.115 |
| Preferred Language: Spanish | 0.168 | 0.179 | 1.18 | (0.83, 1.68) | 0.349 |
| Preferred Language: Chinese | 0.083 | 0.111 | 1.09 | (0.87, 1.35) | 0.453 |
| Preferred Language: Other Non-English | 0.168 | 0.106 | 1.18 | (0.96, 1.46) | 0.112 |
| Clinic Category: Primary Care or Internal Medicine | -0.193 | 0.060 | 0.82 | (0.73, 0.93) | 0.001 |
| HPI Index (per 10 pt) | -0.016 | 0.012 | 0.98 | (0.96, 1.01) | 0.195 |
| **Diastolic BP × Male** | **-0.144** | **0.072** | **0.87** | **(0.75, 1.00)** | **0.045** |

**Supplemental eTable T9. Sex Interaction Coefficient with BMI.**
The final row reports the interaction coefficient, while preceding rows show covariate-adjusted hazard ratios for progression to dementia.

| Term | β (coef) | SE | Hazard Ratio | 95% CI | p-value |
|---|---|---|---|---|---|
| BMI | -0.020 | 0.010 | 0.98 | (0.96, 1.00) | 0.039 |
| Sex: Male | 1.067 | 0.401 | 2.91 | (1.32, 6.38) | 0.008 |
| Type II Diabetes (history) | 0.077 | 0.060 | 1.08 | (0.96, 1.22) | 0.201 |
| Diastolic BP | -0.083 | 0.043 | 0.92 | (0.85, 1.00) | 0.055 |
| Systolic BP | 0.112 | 0.025 | 1.12 | (1.07, 1.17) | <0.001 |
| Age at MCI | 0.055 | 0.004 | 1.06 | (1.05, 1.06) | <0.001 |
| Race/Ethnicity: Other Identity | -0.001 | 0.111 | 1.00 | (0.80, 1.24) | 0.996 |
| Race/Ethnicity: Latinx | 0.131 | 0.121 | 1.14 | (0.90, 1.45) | 0.281 |
| Race/Ethnicity: Asian | -0.028 | 0.083 | 0.97 | (0.83, 1.14) | 0.734 |
| Race/Ethnicity: Black or African American | 0.049 | 0.123 | 1.05 | (0.82, 1.34) | 0.691 |
| Marital Status: Married/Partnered | 0.013 | 0.097 | 1.01 | (0.84, 1.23) | 0.895 |
| Marital Status: Single | -0.247 | 0.116 | 0.78 | (0.62, 0.98) | 0.033 |
| Marital Status: Unknown/Other | -0.417 | 0.365 | 0.66 | (0.32, 1.35) | 0.253 |
| Marital Status: Widowed | 0.144 | 0.103 | 1.16 | (0.94, 1.42) | 0.162 |
| Preferred Language: Spanish | 0.116 | 0.179 | 1.12 | (0.79, 1.60) | 0.516 |
| Preferred Language: Chinese | 0.086 | 0.110 | 1.09 | (0.88, 1.35) | 0.434 |
| Preferred Language: Other Non-English | 0.157 | 0.105 | 1.17 | (0.95, 1.44) | 0.137 |
| Clinic Category: Primary Care or Internal Medicine | -0.188 | 0.060 | 0.83 | (0.74, 0.93) | 0.002 |
| HPI Index (per 10 pt) | -0.014 | 0.012 | 0.99 | (0.96, 1.01) | 0.263 |
| **BMI × Male** | **-0.041** | **0.016** | **0.96** | **(0.93, 0.99)** | **0.008** |

**Supplemental eTable T10. Sex Interaction Coefficient with Type II Diabetes.**
The final row reports the interaction coefficient, while preceding rows show covariate-adjusted hazard ratios for progression to dementia.

| Term | β (coef) | SE | Hazard Ratio | 95% CI | p-value |
|---|---|---|---|---|---|
| Type II Diabetes (history) | 0.099 | 0.078 | 1.10 | (0.95, 1.29) | 0.202 |
| Sex: Male | 0.037 | 0.074 | 1.04 | (0.90, 1.20) | 0.618 |
| BMI | -0.035 | 0.008 | 0.97 | (0.95, 0.98) | <0.001 |
| Diastolic BP | -0.087 | 0.043 | 0.92 | (0.84, 1.00) | 0.045 |
| Systolic BP | 0.113 | 0.025 | 1.12 | (1.07, 1.17) | <0.001 |
| Age at MCI | 0.055 | 0.004 | 1.06 | (1.05, 1.06) | <0.001 |
| Race/Ethnicity: Other Identity | 0.008 | 0.111 | 1.01 | (0.81, 1.25) | 0.940 |
| Race/Ethnicity: Latinx | 0.144 | 0.121 | 1.16 | (0.91, 1.47) | 0.234 |
| Race/Ethnicity: Asian | -0.029 | 0.083 | 0.97 | (0.83, 1.14) | 0.731 |
| Race/Ethnicity: Black or African American | 0.063 | 0.123 | 1.07 | (0.84, 1.36) | 0.607 |
| Marital Status: Married/Partnered | 0.007 | 0.097 | 1.01 | (0.83, 1.22) | 0.941 |
| Marital Status: Single | -0.249 | 0.116 | 0.78 | (0.62, 0.98) | 0.032 |
| Marital Status: Unknown/Other | -0.427 | 0.365 | 0.65 | (0.32, 1.33) | 0.242 |
| Marital Status: Widowed | 0.139 | 0.103 | 1.15 | (0.94, 1.41) | 0.179 |
| Preferred Language: Spanish | 0.110 | 0.179 | 1.12 | (0.79, 1.59) | 0.540 |
| Preferred Language: Chinese | 0.092 | 0.110 | 1.10 | (0.88, 1.36) | 0.407 |
| Preferred Language: Other Non-English | 0.161 | 0.105 | 1.17 | (0.96, 1.44) | 0.126 |
| Clinic Category: Primary Care or Internal Medicine | -0.182 | 0.060 | 0.83 | (0.74, 0.94) | 0.003 |
| HPI Index (per 10 pt) | -0.014 | 0.012 | 0.99 | (0.96, 1.01) | 0.258 |
| **Type II Diabetes (history) × Male** | **-0.051** | **0.114** | **0.95** | **(0.76, 1.19)** | **0.655** |

**Supplemental eTable T11. Model comparison statistics for selecting spline vs. linear specifications for key continuous exposures.**
For each variable, we report the AIC for linear and spline models, the AIC difference (ΔAIC), and the likelihood ratio test (LRT) p-value. Spline models were preferred for systolic and diastolic blood pressure based on both AIC reduction (ΔAIC > 10) and significant LRT results ($p < 0.05$). A spline model was not preferred for BMI.

| Variable | Linear AIC | Spline AIC | ΔAIC | LRT p-value | Spline Preferred? |
|---|---|---|---|---|---|
| **Systolic BP** | 22862.60 | 22849.76 | 12.84 | <0.05 | Yes |
| **Diastolic BP** | 22882.90 | 22862.49 | 20.41 | <0.05 | Yes |
| **BMI** | 22858.85 | 22862.82 | 3.97 | 0.9855 | No |

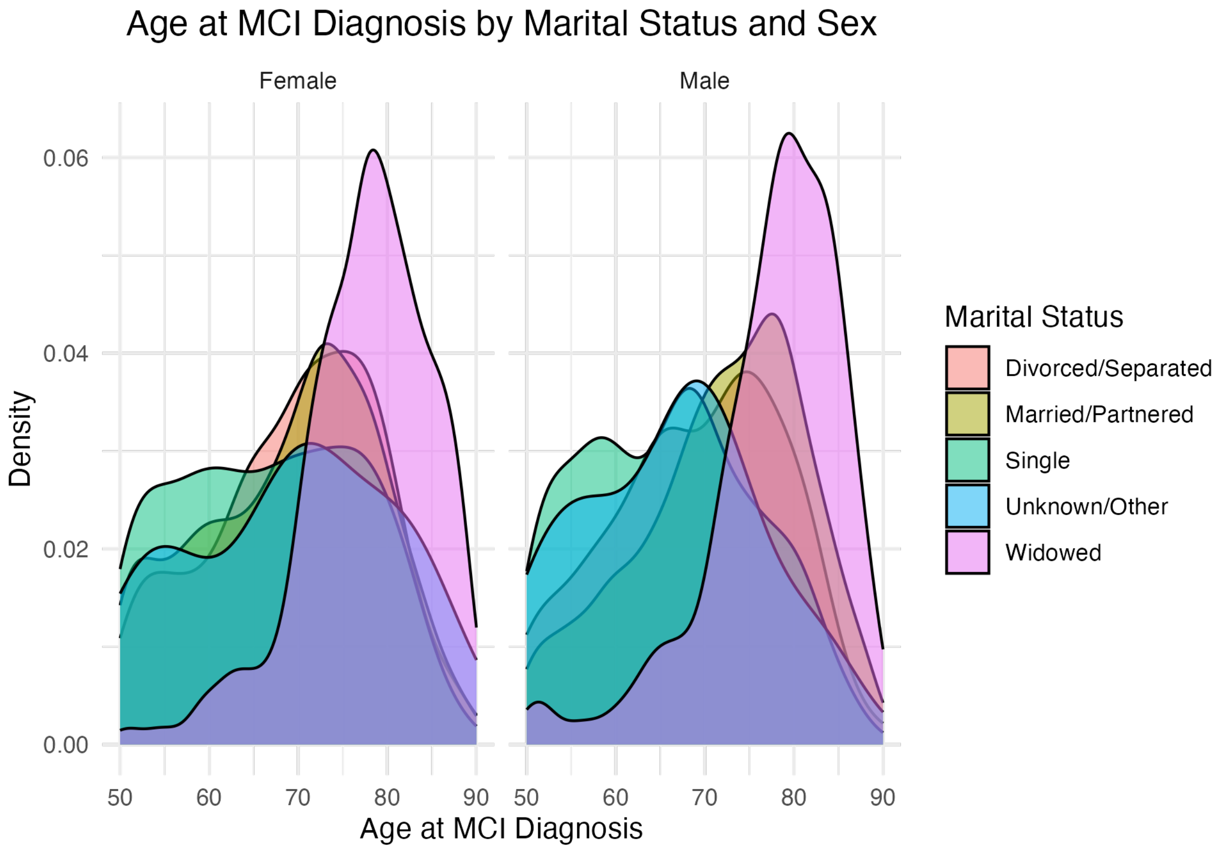


**Supplemental eFigure F1**. **Sex-stratified Age Distributions at Index MCI Diagnosis Across Racial and Ethnic Groups.**
Kernel density estimates are shown for each group, stratified by female (left) and male (right). Densities are scaled within sex strata to illustrate differences in age distribution patterns across racial/ethnic categories.

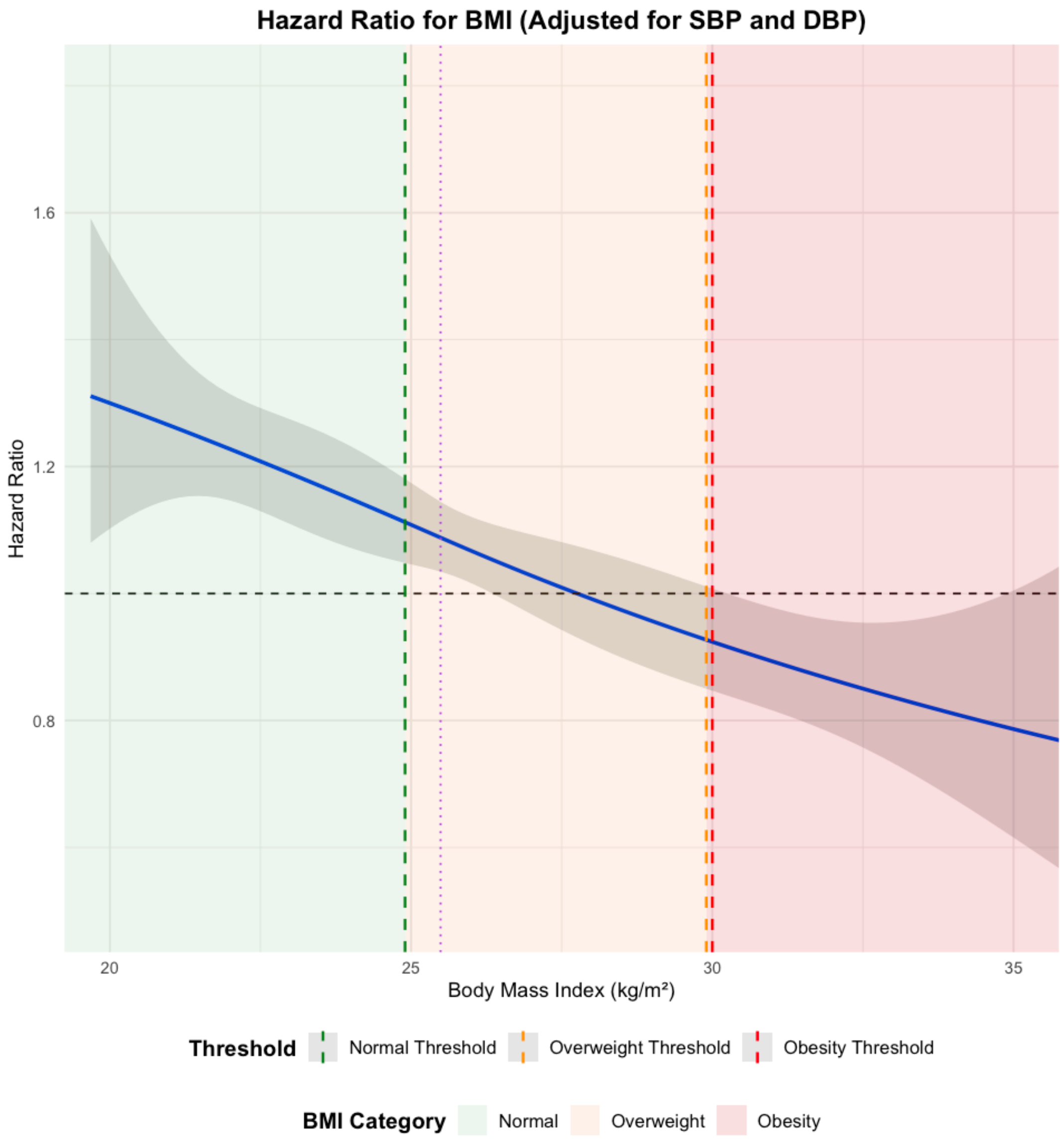


**Supplemental eFigure F2. Spline Model Showing the Hazard Ratio for Progression from MCI to Dementia as a Function of Body Mass Index (BMI), Adjusted for Systolic and Diastolic Blood Pressure.**
The shaded region represents the 95% confidence interval. Vertical dashed lines indicate clinical thresholds for normal weight (green), overweight (orange), and obesity (red), with corresponding shaded background colors for BMI categories.

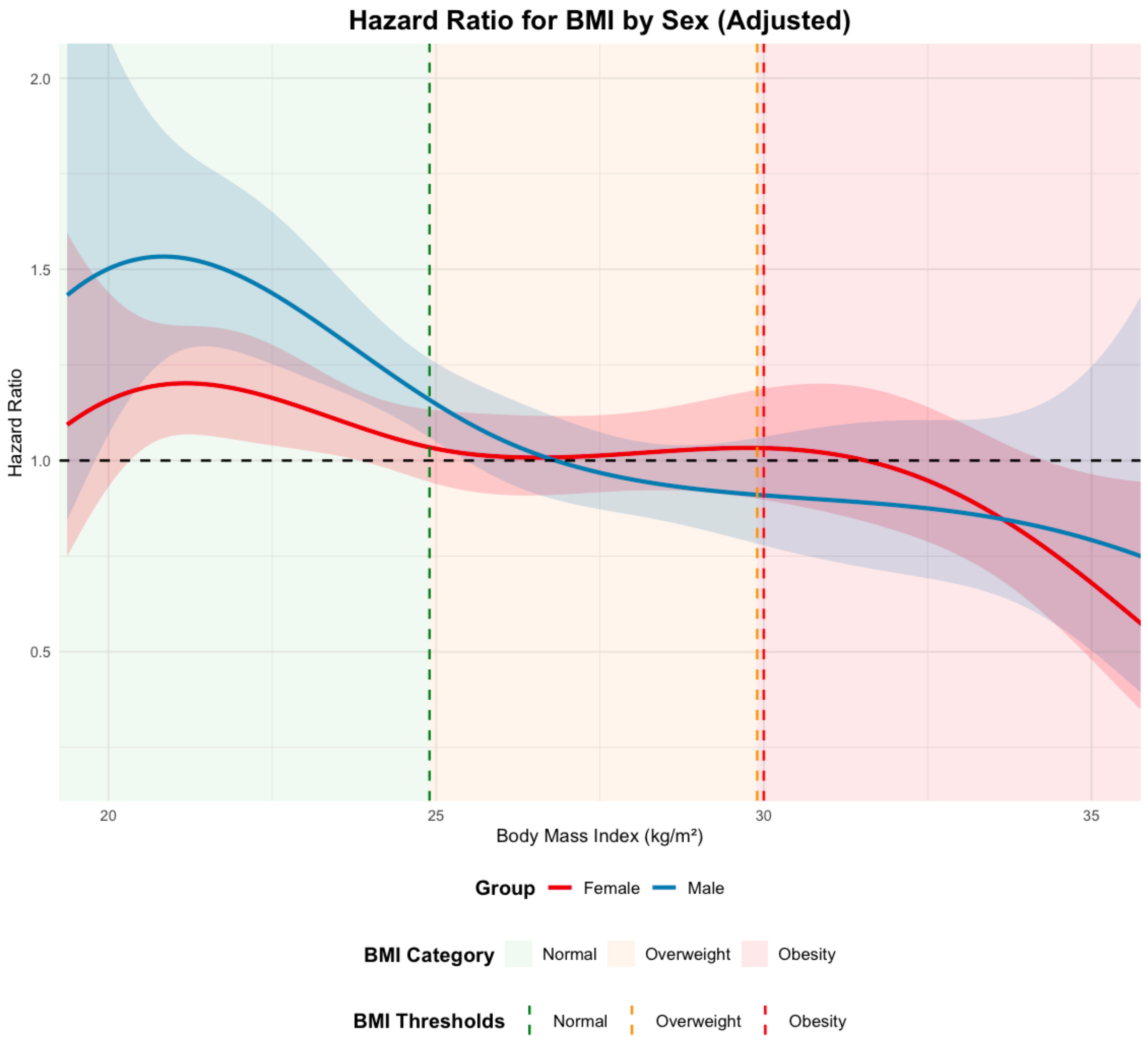


**Supplemental eFigure F3. Sex-Stratified Spline Models Showing the Hazard Ratio for Progression from MCI to Dementia as a Function of Body Mass Index (BMI), Adjusted for Systolic and Diastolic Blood Pressure.**
The shaded regions represent 95% confidence intervals. Vertical dashed lines denote clinical BMI thresholds for normal weight (green), overweight (orange), and obesity (red), with corresponding background shading.
